# Social determination of health and intersectionality in child hunger: A multilevel qualitative comparative case study in Chocó and Risaralda, Colombia

**DOI:** 10.64898/2026.11.19.26368251

**Authors:** Paula Andrea Taborda Restrepo, Yadira Eugenia Borrero Ramírez, Lorena Patricia Mancilla López

## Abstract

Acute child malnutrition has historically been approached in public health as an individual nutritional deficit, despite its persistence in territories marked by structural inequity. This study analyzed how the general, particular, and singular domains of the social determination of health articulate with the intersectional axes of gender, ethnicity, class, and territory in the social production of child hunger in Chocó and Risaralda, two Colombian departments with a high burden of acute child malnutrition. We conducted a qualitative comparative case study grounded in a socio-critical paradigm and a critical feminist epistemology, based on fieldwork carried out in 2024. In-depth interviews with 28 participants—mothers and caregivers, health workers, and decision-makers—were combined with a field diary and a documentary review. Analysis followed open coding, axial coding, and categorical integration, yielding 722 coded units and 207 analytical categories, which were organized through a convergence matrix across the three domains of social determination and the intersectional axes. In the particular domain, informal work, food restriction, and institutional fragmentation shaped ways of life marked by the feminization of care, while formally established care pathways failed to sustain nutritional recovery. In the singular domain, trajectories of accumulated violence, forced early unions, and female genital mutilation preceded and shaped motherhood under precarious conditions, together with guilt, anguish, and fear produced by the institutional moralization of maternal care. Across domains, structural conditions were subsumed into collective ways of life and embodied in singular trajectories, without cancelling situated forms of agency, resistance, and care. A multilevel integration of social determination and intersectionality allows child hunger to be understood as a historically produced process that simultaneously traverses structures, ways of life, and bodies, and that cannot be resolved through exclusively clinical or welfare-based responses. The proposed model offers a transferable analytical tool for critical public health research seeking to integrate structural, collective, and biographical evidence.

## Introduction

Acute child malnutrition has historically been addressed by public health as an individual problem of nutritional deficit, correctable through anthropometric surveillance and food supplementation. However, in Colombian departments such as Chocó and Risaralda, where child mortality associated with malnutrition remains a persistent burden despite the normative and programmatic strengthening of recent decades, this individualizing approach shows clear explanatory limits. In response, the social determination of health—a perspective developed within Latin American collective health—proposes understanding health–disease processes as the result of the way society organizes the production, distribution, and reproduction of life, rather than as the sum of isolated risk factors [1]. From this perspective, child hunger ceases to be a biomedical event confined to the child’s body and becomes the situated expression of historical power relations that unequally distribute the possibilities of living, falling ill, and dying [2].

The analytical architecture of social determination organizes these processes into three dialectically articulated domains: general, particular, and singular. The general domain corresponds to historically produced structural conditions—structural racism, territorial dispossession, armed conflict, persistent poverty—that constitute the conditions of production of the phenomenon rather than its external context. The particular domain refers to the ways of life of collectives: patterns of work, consumption, care, and relationship with nature shaped by class, gender, and ethnicity. The singular domain corresponds to the space where these conditions are embodied in biographical trajectories, specific bodies, and lived experiences [3]. These domains operate neither independently nor hierarchically: they are articulated through subsumption, whereby the logics of the general domain condition the possibilities of the particular and singular domains, and through relative autonomy, which recognizes situated margins of action, resistance, and transformation [3].

A frequent use of this framework, however, tends to privilege analysis of the general domain—usually through aggregate epidemiological indicators—without explicitly operationalizing how these conditions translate into concrete ways of life and become embodied in singular experiences. This limitation is particularly relevant when the framework is articulated with intersectionality, which makes it possible to understand how structural inequities differentially affect people according to their specific position at the intersection of gender, ethnicity, class, and territory, avoiding treating these categories as independent variables or as a simple sum of disadvantages [4,5]. The articulation between social determination and intersectionality has been identified as necessary to refine the analysis of health inequities, but its empirical operationalization in field research remains scarcely documented in the Latin American public health literature [3,6].

This study sought to address this gap by operationalizing an analytical model that explicitly articulates the three domains of social determination with the intersectional axes of gender, ethnicity, class, and territory, applied to understanding child hunger in two Colombian departments with contrasting structural configurations: Chocó, characterized by generalized structural deprivation, and Risaralda, where exclusion is concentrated in specific ethnic-territorial enclaves. The epidemiological component of the general domain in these territories is reported in a separate manuscript; this article focuses on the particular and singular domains and their articulation with the general domain, with the aim of showing how structural inequities translate into concrete ways of life and become embodied in the biographical trajectories of women caregivers, as narrated by them and by health workers and decision-makers. The aim of this article was therefore to analyze the articulation between the general, particular, and singular domains of the social determination of health and the intersectionality of gender, ethnicity, class, and territory in the social production of child hunger in Chocó and Risaralda.

## Materials and methods

### Study design and setting

We conducted a qualitative comparative case study [7], framed within a socio-critical paradigm and a critical feminist epistemology, as part of a broader doctoral research project on the social determination of acute child malnutrition in Chocó and Risaralda, Colombia. The comparative case design pursued analytical rather than statistical generalization, in the sense proposed by Yin, which made it possible to identify patterns and configurations that transcend the particular cases [7]. The two departments were selected through purposive theoretical sampling aimed at maximizing structural contrast: Chocó, with historical conditions of exclusion and generalized structural poverty, and Risaralda, with better aggregate departmental indicators that nonetheless conceal critical pockets of ethnic-territorial exclusion in municipalities with Emberá Chamí Indigenous populations. Fieldwork was carried out in 2024 in both departments and included nutritional recovery centers and rural communities.

### Participants and sampling

Twenty-eight participants were included: mothers and caregivers of children under five years of age treated for moderate or severe acute malnutrition in the previous two years; health care personnel with at least one year of experience in the care of affected children; and institutional actors with decision-making capacity in epidemiological surveillance, policy formulation, or program implementation. A caregiver was defined as the person who devotes most of their time to the child’s care. Sampling was purposive, aimed at maximizing the diversity of experiences and positions relevant to the study aims. Caregivers were contacted at the nutritional recovery centers in the study territories, and all people invited agreed to participate. Participants were recruited between 1 July 2024 and 30 September 2024. Participants were distributed between Chocó (n = 16) and Risaralda (n = 12) and comprised caregivers (n = 8, all adult women, ranging in age from 18 to over 50 years), nutritionists (n = 4), a nurse (n = 1), a physician (n = 1), program coordinators (n = 3), a social worker (n = 1), decision-makers in public and institutional management (n = 8), and traditional midwives or breastfeeding counselors (n = 2). People who did not provide informed consent and caregivers of children whose malnutrition was associated with specific secondary clinical conditions—cerebral palsy, severe dysphagia, or esophageal atresia—were excluded, in order to keep the focus on malnutrition linked to social determinants.

### Data collection

Data were collected through semi-structured in-depth interviews guided by thematic guides that prioritized biographical accounts of the child’s malnutrition, household living conditions, and experiences of institutional care [8]. Two types of guide were used: a thematic biographical interview for caregivers and a semi-structured interview for health professionals and decision-makers; both were applied flexibly, following the dynamics of each conversation. The guides were pilot tested before fieldwork, and each participant was interviewed once. Interviews were conducted by the first author, a nutritionist-dietitian and doctoral candidate in public health with prior professional experience in the treatment of severe acute malnutrition and in the epidemiological surveillance of malnutrition deaths, who adopted a reflexive and relational stance toward participants [8]; they lasted approximately one hour on average and took place in locations agreed with participants that ensured privacy. With the exception of one decision-maker in Chocó who was previously known to the interviewer, there was no prior relationship with participants. Interviews with Emberá participants were conducted with the support of local translators. All interviews were audio-recorded with informed consent, transcribed verbatim—including pauses, silences, and relevant emotional expressions—by trained external staff under confidentiality agreements, and reviewed and anonymized by the first author before analysis; transcripts were not returned to participants. In addition, the first author moderated a focus group lasting approximately two hours with eight hospital workers involved in the care of children with acute malnutrition, in order to explore collective institutional perspectives; quotations from this source are identified as “focus group”. A field diary was kept with contextual observations, emerging analytical notes, and reflections on the researcher’s position, including the territorial security conditions that shaped fieldwork, among them an armed strike that delayed access to one of the study sites. A documentary review of public policies, technical guidelines, constitutional case law related to the right to food, and institutional documents from both departments was also carried out, with a contextual and interpretive function rather than as an independent component of the analysis.

### Data analysis

Qualitative analysis was conducted through coding in three phases: open coding to identify units of meaning, axial coding to establish relationships between categories, and categorical integration to articulate interpretive configurations across the general, particular, and singular domains of social determination [9]. Analysis was carried out in parallel with fieldwork, which allowed data collection to be adjusted to explore emerging themes; data collection ended when new interviews no longer yielded new categories (data saturation). The process generated 722 coded units and 207 analytical categories, organized in a Microsoft Excel matrix that facilitated the systematization and comparison of patterns across actors and territories without replacing the interpretive process. Analysis was conducted by the first author and discussed with the co-authors throughout the process. Final integration was performed through a convergence matrix organized by objective, territory, and domain of determination, which made it possible to examine convergences, complementarities, and divergences between documentary sources and interviews, preventing one component from functioning merely as an illustration of another. Epidemiological findings for the general domain, reported in a separate manuscript, were incorporated into this analysis only as a contextual reference for interpreting their articulation with the particular and singular domains. Quotations are identified by anonymized participant codes indicating actor type (M, mother or caregiver; Abuela, grandmother caregiver; Trab, health worker; Tom, decision-maker) and department (Ch, Chocó; R, Risaralda); they were translated from Spanish into English by the authors. Once analysis was completed, results were returned to participants through a narrated story, built from the accounts of the different actors and shared as a video, which was sent to all participants together with a summary of the results.

### Ethics statement

The study complied with Colombian Resolution 8430 of 1993 [10] and was approved by the Research Ethics Committee of the Facultad Nacional de Salud Pública, Universidad de Antioquia (certificate 21030002-003-2024), which classified it as greater-than-minimal risk and issued specific recommendations for research with Indigenous peoples, including the condition that fieldwork could not begin until the informed consent form had been translated into the Emberá language. Access to communities was preceded by processes of socialization and agreement with local, Indigenous, and ethnic authorities; in Emberá communities, oral authorization was granted by the Indigenous governor. Written informed consent was obtained from all participants, using forms in Spanish and in the Emberá language, with the support of local translators when necessary. All participants were adults (18 years of age or older), and none received financial compensation for their participation. Psychological support, following a psychological first aid protocol, was available in the event of emotional distress arising from the narration of painful experiences, and all participants were anonymized through codes. To protect participants’ confidentiality, ages, precise locations, family details, and other indirectly identifying information have been removed or generalized in this article; interview transcripts and detailed biographical data are not publicly available, and readers may contact the corresponding author to request access to these data, subject to approval by the Research Ethics Committee. The study followed the International Ethical Guidelines for Health-related Research Involving Humans of the Council for International Organizations of Medical Sciences [11] and the principles of the Declaration of Helsinki [12]. The study is reported in accordance with the Consolidated Criteria for Reporting Qualitative Research (COREQ) [13] (S1 Checklist). Additional information regarding the ethical, cultural, and scientific considerations specific to inclusivity in global research is included in the Supporting information (S2 Checklist).

## Results

The analytical model articulating social determination and intersectionality developed for this study is summarized in Fig 1, which synthesizes the processes observed in each domain, the relationship of subsumption between them, and the cross-cutting axis of gender, racialization, and moralization of care that traverses them.

**Fig 1.**
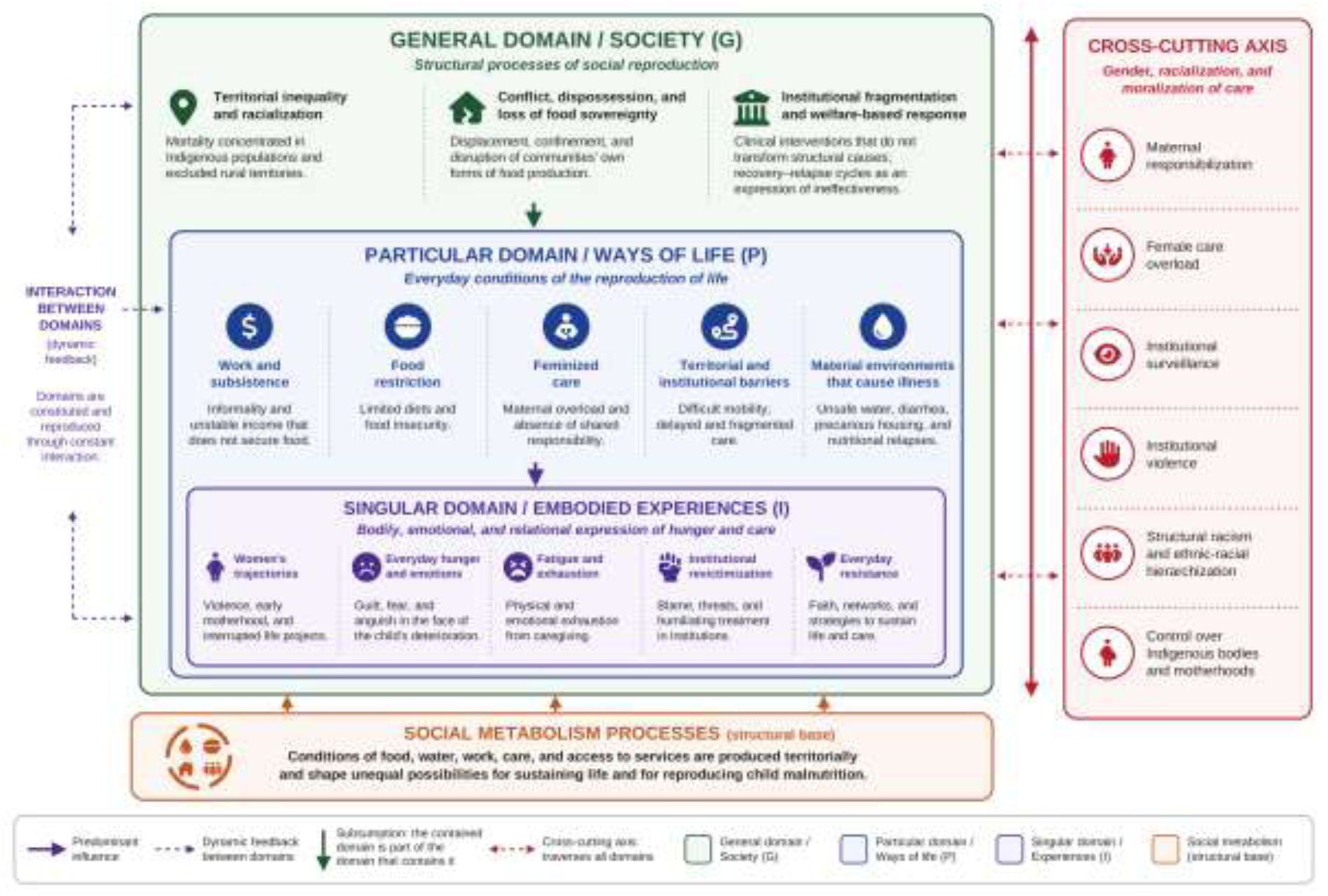
Social determination of child hunger: articulation between domains and the cross-cutting axis of gender and racialization. Authors’ elaboration, adapted from Breilh [3]. Icons from Font Awesome Free (https://fontawesome.com), licensed under CC BY 4.0.

### Particular domain: ways of life shaped by the feminization of care

The structural conditions of the general domain do not act directly on children’s bodies: they are first translated into concrete ways of life that organize work, consumption, care, and relationships with institutions within households and communities [3].

### Work and food consumption

In both departments, household income did not constitute a stable base from which to plan food, but rather a daily variable organized around urgency. Accounts described fragmented income from occasional informal work (”below the minimum wage”, M-Ch-1) or from informal work combined with occasional sales (M-R-1). When daily income was insufficient, households reorganized the frequency, quantity, and quality of meals: one mother described her child’s daily diet as “plantain, rice and, when there is some, a little panela [unrefined cane sugar]” (M-R-1); another noted that she sometimes merged breakfast and lunch because there was not always food (M-Ch-2). These strategies of reducing and merging meals do not reflect a lack of nutritional knowledge: when one mother was asked what she would like to receive as food assistance, she answered “vegetables, fruit, milk, things like that” (M-Ch-1), revealing the gap between knowing, wanting, and being able to access food [14]. The same precariousness permeated the institutional response: a decision-maker in Chocó described epidemiological surveillance sustained by contractors hired late and intermittently—”contracts start late…deaths are frequent” (Tom-Ch-2)—showing that informality not only affects families but is reproduced within the very state apparatus responsible for responding to the problem.

### Organization of care

In both territories, the organization of child care followed a consistent pattern: women assumed full responsibility for children’s feeding, supervision, and well-being, a pattern legitimized even by the women themselves: “as women we are specialized in that” (M-Ch-1). A health professional confirmed this from their territorial experience: “the head of a household is the woman…the mother is the one who mostly has the responsibility” (Trab-Ch-3). Male control also extended to economic resources intended for care: “the father is waiting at the door for her to hand him the money [from the subsidy]” (health worker focus group), which may reduce the protective effect of conditional cash transfers aimed at children.

### Institutional response and cultural mediations

Care pathways exist formally, but, as one decision-maker summarized, “the pathway exists, but it does not walk” (Tom-Ch-2). Post-discharge follow-up revealed the clearest limit of the model: “when those children go back home, they return to the same situations…that ends up becoming a vicious circle for us” (Tom-Ch-1), showing that the system intervenes correctly on the individual body without being able to modify the conditions to which the child returns. In Emberá communities, malnutrition was not uniformly classified as a biomedical entity: the Jaibaná (traditional Emberá healer) acts as a therapeutic authority who intervenes in the decision to seek care in the health system—”until the Jaibaná says: this is not mine…you can take him” (Tom-Ch-2)—within a culturally structured, non-arbitrary system of medical pluralism [15,16]. One mother attributed her child’s deterioration to malaria rather than malnutrition, an interpretation consistent with a context where this disease is endemic and where the biomedical category of malnutrition may have no direct equivalent in the Emberá classification system (M-R-1). The institutional response showed limited capacity for dialogue with these systems: technologies delivered without appropriation—”they have built them smokeless stoves…and they are cooking on stones” (focus group)—and donated foods that go unprepared because they are incompatible with the available domestic conditions.

### Singular domain: trajectories, emotions, and agency in precarious motherhood

The singular domain showed how general conditions and collective ways of life are embodied in the concrete trajectories, bodies, and experiences of women caregivers [3,17].

### Trajectories of accumulated violence

One young mother, when asked about the most difficult moments of her life before becoming a mother, reconstructed a chain of structural ruptures that began in early childhood: sexual abuse, family disbelief (”but she didn’t believe me”, M-Ch-5), separation from her family of origin, a legal complaint that did not prosper (”they told me no…that there were no witnesses”, M-Ch-5), and an adolescent pregnancy that interrupted her schooling, years before her own child required nutritional recovery. When asked whether she still felt resentment, she replied: “I still feel resentment” (M-Ch-5). This trajectory cannot be read as an exceptional story of individual misfortune: the child’s malnutrition does not begin at birth, but much earlier, in the mother’s unprotected childhood [18]. Fig 2 synthesizes this trajectory, together with the accounts of other women caregivers and the field diary records, as a composite life course in which successive ruptures accumulate before the child’s malnutrition becomes clinically visible.

**Fig 2.**
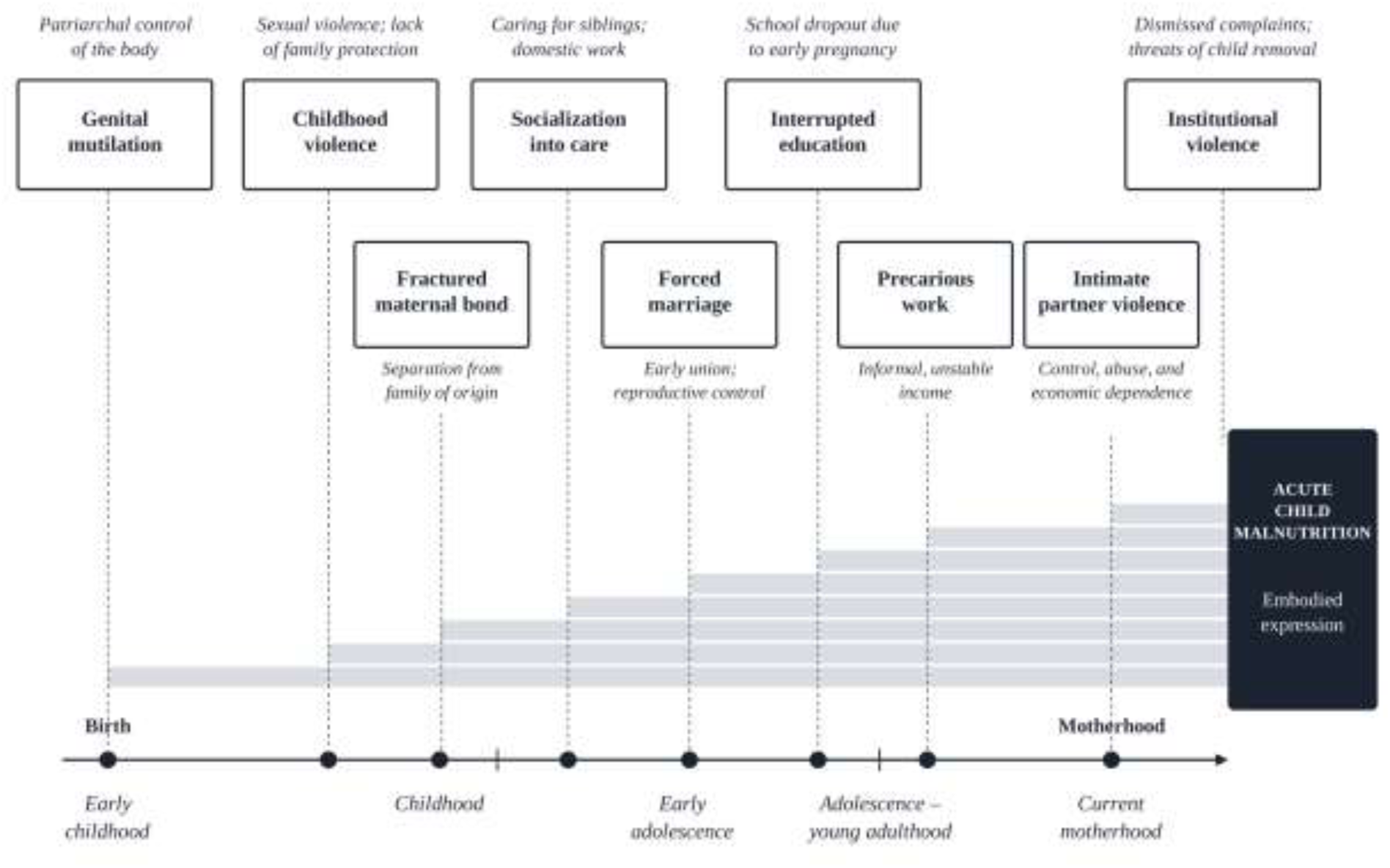
Life trajectories and the reproduction of precariousness in the singular domain. Composite trajectory reconstructed by the authors from the in-depth interviews with women caregivers and the field diary (2024); it does not represent a single participant, and ages and other potentially identifying details have been omitted.

A narrative with a similar structural weight came from another mother, interviewed through a translator, who had been forced into a union during childhood [19] and was raising her children alone. She described being unable to make decisions about her own reproduction (”she did not use family planning…the husband did not want to”, M-R-1) and a structural loneliness sustained over time: “she is always alone with the children, she feels weary with them” (M-R-1). In addition, the field diary recorded—although it was not the subject of the structured interviews—the silenced persistence of female genital mutilation among Emberá girls in the region. In Colombia, this practice has been documented mainly in Emberá communities of Risaralda and Chocó: 204 cases were reported between 2020 and 2025, 177 of them in Indigenous girls and more than half in girls aged zero to five years; cases reach the health system predominantly through complications such as sepsis, which also indicates considerable underreporting [20]. Emberá community leadership itself has promoted its eradication since 2010, a process that in 2026 led to Law 2609, the first specific legislation in Latin America prohibiting the practice and mandating a public policy for its prevention, care, and eradication with an intercultural approach [21]. The co-occurrence of forced early union, motherhood without reproductive autonomy, and genital mutilation in the same territory allows these processes to be read as a single inscription of patriarchal control over girls’ bodies, which precedes and prefigures the malnutrition of their future children.

### Emotions: institutionally produced guilt, anguish, and fear

The emotions running through the narratives were not random individual reactions but socially produced processes [22]. Anguish in the face of diagnostic uncertainty emerged most persistently: “it distresses me a lot…a thousand things go through my head” (M-Ch-1). Maternal guilt was the most cross-cutting emotional process: “yes, I feel guilty, because financially…there were no…resources” (M-Ch-1); another mother, who had stopped breastfeeding to care for a seriously ill relative, wondered: “if I had been there…I think he would not have lost weight” (M-Ch-4). In no case did guilt correspond to an objective failure of care, but rather to structural conditions—insufficient income, the impossibility of being with two sick relatives at the same time, the child’s previous illnesses—reinterpreted by the mothers themselves as personal failure, which can be read as a form of symbolic violence [23]. This guilt did not emerge spontaneously: one mother reported that, after missing a check-up to care for another of her children, institutional staff warned her that her daughter could be taken away (”that if something happens to the girl they will take her away from me”, M-Ch-1), producing a fear that, as a health worker in Risaralda confirmed, keeps other families away from the system that could care for them (”they don’t go to the hospital…I don’t know whether to take him”, Trab-R-4).

### Agency and sustained care

The narratives were not limited to suffering. Asked about the happiest moment of her life, the mother who had been abused in childhood answered without hesitation by naming her children (M-Ch-5); another framed happiness as the very survival of her children: “knowing that they are alive, that they are well…there we go” (M-Ch-1). Faith operated as a genuine protective resource: “I believe in God…I paid attention to his feeding…and he recovered” (M-Ch-1). And the demand for food autonomy expressed by a caregiver in Risaralda—”support is needed to grow crops in the resguardo [Indigenous reserve]…to have their own crops to draw from” (Abuela-R-2)—did not express a request for charity but the affirmation of a right to produce one’s own food [24]. These forms of agency, resistance, and situated knowledge do not negate the conditions of oppression described; they show that, even in precariousness, women caregivers deploy active strategies to sustain their children’s lives [25,26].

### Articulation between domains: subsumption and relative autonomy

The joint reading of the three domains revealed the two dialectical movements that social determination attributes to its analytical architecture: subsumption and relative autonomy [3]. Subsumption became visible in the trajectory of M-Ch-5: the structural deprivation of the general domain—poverty and the institutional fragmentation of child protection—was translated into a particular way of life marked by the absence of family protection networks, and was embodied in the singular domain as sexual abuse that was not believed, being handed over to third parties, and adolescent motherhood without reproductive autonomy. Similarly, the loss of food sovereignty in the general domain was translated in the particular domain into household strategies of reducing and merging meals, and was embodied in the singular domain in the anguish and guilt of mothers who knew, without being able to guarantee it, the diet that would have better protected their children.

At the same time, relative autonomy became visible in the forms of agency identified across domains: in the particular domain, in the persistence of Emberá knowledge systems that guide therapeutic decisions according to their own logic; in the singular domain, in faith, filial love, and the explicit demands for food sovereignty voiced by women caregivers. None of these expressions reverses the structural conditions that produce them, but they show that individuals and collectives have situated margins of action and resistance that analysis cannot omit without reducing families to passive victims [3]. This articulation also explains why the cross-cutting axis of gender and structural racism traverses the three domains with different densities: it is expressed in the general domain as the territorial and ethnic-racial concentration of risk, in the particular domain as the feminization of care work and the male capture of resources intended for children, and in the singular domain as an institutional moralization that shifts onto women responsibilities that exceed their material possibilities.

## Discussion

The main contribution of this article is the empirical operationalization of the articulation between social determination and intersectionality through a three-domain model that does not remain at the general level of aggregate indicators, but explicitly shows how structural inequities are translated into collective ways of life and embodied in singular biographical trajectories. This exercise responds to a recurrent critique of the social determinants of health approach: its tendency to fragment reality into isolated variables and to propose interventions that do not question the structures that produce inequality [1]. By showing the subsumption of structural processes into concrete bodily experiences, the model proposed here provides empirical support for the argument that child hunger cannot be understood or addressed solely through the biomedical management of the nutritional event.

The findings of the singular domain constitute, in particular, a little-documented contribution to the literature on child malnutrition in Colombia: the identification of a chain in which childhood sexual violence, forced early union, and female genital mutilation precede and shape motherhood under conditions of extreme precariousness. This finding is consistent with evidence on historically structured biographical trajectories, according to which inequality accumulates and settles at different moments of the life course [27], and suggests that a child’s malnutrition does not begin at the moment of clinical diagnosis but in processes that started much earlier in the mother’s life [18]. Although female genital mutilation, recorded in the field diary, was not directly addressed in the interviews, it cannot be treated as anecdotal: recent Colombian legislation prohibiting the practice and mandating its care with an intercultural approach [21] shows that it is a recognized public health and human rights problem beyond this study, and reinforces the need for child malnutrition surveillance systems to incorporate indicators of gender-based violence in childhood.

The analysis of the particular domain showed that the institutional moralization of maternal care—expressed in threats to remove custody when check-ups are missed because of precariousness itself—constitutes a mechanism through which the health system shifts onto women structural responsibilities that it cannot itself guarantee. This reading is consistent with the notion of a crisis of care, according to which economies depend on women’s reproductive work without guaranteeing the conditions for its sustainability [28], and with the characterization of maternal guilt as a form of symbolic violence in which the consequences of structural conditions come to be perceived as personal failure [23]. Far from being a side effect of care, our findings suggest that this moralization operates as an active barrier to access: mothers and health workers agreed that the fear of losing custody of a child keeps families away from the system that could care for them.

The medical pluralism identified in Emberá communities in Risaralda—the mediation of the Jaibaná and the classification of malnutrition as malaria within their own knowledge system—should not be interpreted as a cultural barrier to be overcome through health education, but as a culturally structured system of diagnosis and care with which the biomedical system has failed to engage in effective dialogue [15,16]. Attributing delays in care-seeking exclusively to “cultural beliefs”—as some of the institutional staff interviewed did—shifts responsibility onto families and obscures the geographic, linguistic, and institutional-trust barriers that actually shape access [29].

From a public health perspective, these findings suggest three priority lines of action. First, incorporating routine screening for gender-based violence and early unions into care protocols for acute child malnutrition, in coordination with child protection pathways. Second, replacing custody threats with social support strategies that recognize families’ material barriers, in line with the reinforced protection recognized by the Colombian Constitutional Court regarding timely, continuous, and comprehensive access to health care [30] and minimum guarantees of water and sanitation in Indigenous territories [31]. Third, building intercultural care models that incorporate, under conditions of horizontal dialogue rather than subordination, the knowledge systems and therapeutic authorities of the Emberá people, instead of treating them as obstacles to be overcome through one-way health education.

This study has limitations that should be considered when interpreting the findings. The aggregate epidemiological component of social determination in these territories is reported in a separate manuscript; therefore, this article does not include its own quantitative analysis of the general domain but draws on it as a contextual reference. The qualitative sample size, although adequate for the comprehensive purpose of the study, does not allow statistical generalization; its value lies in the analytical generalization of the proposed model [7]. The study centered the narratives of mothers and caregivers, which entailed lower representation of male voices and of higher-level institutional decision-makers. Although interviews with Emberá mothers were conducted with translation, rendering culturally situated experiences into academic categories entails losses of meaning. Fieldwork was also constrained by territorial security conditions, including an armed strike that delayed access to one of the sites. Finally, female genital mutilation emerged from the field diary rather than from the interview guides, which limits the depth of its documentation in this study and points to the need for specific, culturally appropriate research on the topic, developed in direct dialogue with the Emberá authorities and women’s organizations that have led its eradication.

### Conclusions

The multilevel integration of the social determination of health and intersectionality makes it possible to understand child hunger in Chocó and Risaralda as a historically produced process that simultaneously traverses structures, collective ways of life, and singular bodily trajectories, without cancelling the situated forms of agency and resistance through which women caregivers sustain their children’s lives. Institutional responses centered on individual clinical recovery are necessary but insufficient unless they are articulated with transformations in working conditions, intercultural barriers to access, effective protection against gender-based violence in childhood, and the social redistribution of care. The three-domain model proposed here offers a transferable analytical tool for future critical public health research seeking to integrate structural, collective, and biographical evidence in understanding other expressions of the social determination of health in contexts of high territorial and ethnic inequity.

## Data Availability

Data cannot be shared publicly because of their qualitative and sensitive nature (interviews containing accounts of violence and personal information of women caregivers and Indigenous communities), in accordance with the confidentiality commitments made to participants and the conditions of the ethics approval (certificate 21030002-003-2024, Research Ethics Committee, Facultad Nacional de Salud Publica, Universidad de Antioquia). Data are available upon reasonable, justified request to the corresponding author, subject to authorization by that committee.

## Acknowledgments

This article is derived from the doctoral thesis “’In the cradle of hunger my child was’: understanding the social determination of acute child malnutrition in Chocó and Risaralda in 2024, a comparative case study”, submitted to the Doctorate in Public Health of the Universidad de Antioquia (Facultad Nacional de Salud Pública Héctor Abad Gómez), Epidemiology Research Group, Health Equity research line. We thank the mothers, caregivers, community leaders, health personnel, and institutional actors who shared their experiences, and the local translators who made the interviews with Emberá participants possible.

## Supporting information

**S1 Checklist. COREQ checklist.** Consolidated criteria for reporting qualitative research checklist completed for this study.

**S2 Checklist. Inclusivity in global research questionnaire.**

